# Tracking the prevalence and emergence of SARS CoV2 variants of concern using a regional genomic surveillance program

**DOI:** 10.1101/2023.05.08.23289687

**Authors:** Ana Jung, Lindsay Droit, Binita Febles, Catarina Fronick, Lisa Cook, Scott A. Handley, Bijal A Parikh, David Wang

## Abstract

SARS-CoV-2 molecular testing coupled with whole genome sequencing is instrumental for real-time genomic surveillance. Genomic surveillance is critical for monitoring the spread of variants of concern (VOC) as well as novel variant discovery. Since the beginning of the pandemic millions of SARS-CoV-2 genomes have been deposited into public sequence databases. This is the result of efforts of both national and regional diagnostic laboratories. Here we describe the results of SARS-CoV-2 genomic surveillance from February 2021 to June 2022 at a metropolitan hospital in the USA. We demonstrate that consistent daily sampling is sufficient to track the regional prevalence and emergence of VOC. Similar sampling efforts should be considered a viable option for local SARS-CoV-2 genomic surveillance at other regional laboratories.

## INTRODUCTION

SARS-CoV-2 first appeared in Wuhan, China in late 2019 and was declared a global pandemic by the World Health Organization (WHO) on 11th of March, 2020 [1]. The first SARS-CoV-2 genome sequence was determined in January of 2020 [2]. Since this time over 15 million SARS-CoV-2 genomes have been sequenced and made publicly available [3][4]. This global genomic surveillance project has been an effective way to identify and track nucleotide changes with the potential to influence viral transmission dynamics, pathogenicity, diagnostic performance, vaccine efficacy and immune escape [2,5,6].

Genomic surveillance has enabled classification of emergent SARS-CoV-2 into variants of concern (VOC), variants of interest (VOI), variants being monitoring (VBM), and variants of high consequence (VOHC). This classification is based on their predicted transmissibility, virulence, and ability to cause severe disease. Classification of SARS-CoV-2 variants is changing overtime. Previously classified SARS-CoV-2 VOCs include Alpha (B.1.1.7), Beta (B.1.351), Gamma (P.1), Delta (B.1.617.2), and Omicron (B.1.1.529); VOI included Lambda (C.37) and Mu (B.1.621); and VBM include AZ.5, C.1.2, B.1.617.1*, B.1.526*, B.1.525*, and B.1.630, B.1.640 [7]. As of December 1, 2022, the only VOC lineage is Omicron with Omicron XBB.1.5 being the only VOI as of March 15, 2023.

Information about which VOC are circulating within a regional population is important for public health preparedness and response. In addition, regional genomic surveillance can lead to the original identification of many important VOC. This includes the original detection of Alpha, Beta, Gamma, Delta and Omicron which were first detected in the United Kingdom, South Africa, Brazil, India and multiple countries, respectively [8]. Implementing a regional genomic surveillance program requires significant expense, time and access to modern sequencing technology and bioinformatic expertise. Thoughtful sample selection (size and frequency) is critical to creating a regional genomic surveillance program capable of detecting circulating VOC and novel variant discovery within the confines of local resources.

Here we report the results of a regional SARS-CoV-2 genomic surveillance program run at a metropolitan hospital in St. Louis, MO, USA at a sampling of ∼5 samples/1,000,000 people/week (n = 1,240). Our findings provide evidence that this sampling rate is sufficient to detect the prevalence of known and novel VOC within a community. These results should serve as a guideline for future SARS-CoV-2 genomic surveillance programs.

## MATERIALS AND METHODS

### Ethical considerations

Ethical approval for this study was obtained from the Washington University School of Medicine Institutional Review Board (IRB202004259) with a waiver of consent.

### Sample collection

Nasopharygeal (NP) swab specimens were collected in universal transport medium and submitted for routine clinical SARS-CoV-2 testing at the Barnes-Jewish Hospital Molecular Infectious Disease Lab. Specimens were tested for the presence of SARS-CoV-2 on the cobas SARS-CoV-2 or cobas SARS-CoV-2 & influenza A/B assays performed on the cobas 6800 instrument (Roche) according to manufacturer instructions. Samples positive for SARS-CoV-2 with a minimum cycle threshold (Ct) of 27 for either of two assay targets were eligible for genomic sequencing. 3 to 4 random specimens per day, from the pool of all eligible specimens with sufficient residual volume, were ultimately selected for archival and subsequent sequence analysis. Samples were archived at -70°C in cryovials between 3 and 7 days post-collection.

### SARS-CoV-2 genome sequencing

Total nucleic acid was extracted on a MagNa Pure instrument (Roche) according to the manufacturer recommendations. cDNA was prepared using the ARTIC v3 protocol for samples collected between the dates of February and October, 2021 and the ARTIC v4 protocol between the dates of November, 2021 and June, 2022 [9,10]. The cDNA was purified by a 1x AMPure bead cleanup with a final elution in 10mM TrisHCl, ph 8.5. Purified cDNA was quantitated by a Qubit 1x dsDNA HS Assay (Thermofisher). 50-100ng of the 400bp cDNA amplicons are converted into Illumina libraries on the Ep5075 (Eppendorf) using the KAPA Hyper library prep kit (Roche Diagnostics) using 1/4 of vendor recommended reagents and full length dual indexed adaptors diluted to 250nM [11]. Final libraries were checked for quality and quantity on the LabChipGX instrument (Perkin Elmer), using the DNA High-sensitivity kit. Libraries were normalized to 5nM and an equal volume was pooled per library. This final library pool was quantitated by qPCR using the KAPA Library Quantification kit (Roche Diagnostics) and diluted to 2nM for sequencing in 10mM TrisHCl, pH 8.5. Libraries were loaded at 12pM with a 20% phiX spike in on the MiSeq v3, 600 cycle kit according to Illumina’s guidelines, generating 2×250 reads.

### Analysis of SARS-CoV-2 genomic sequence

We implemented a SARS-CoV-2 genome analysis pipeline that started with raw sequence data and generated quality control information, consensus genomes using the Chan Zuckerberg Biohub SARS-CoV-2 Illumina Pipeline (https://github.com/czbiohub/sc2-illumina-pipeline). Consensus genome lineage assignments were created using both Nextclade (v.2.9.1) and Pangolin (v.4.1.3) [12,13]. Per run phylogenetic trees were generated using Augur and visualized in Microreact [14,15].

### Data Availability

Whole viral genomes were made publicly available at GenBank (**Supplemental Table 1**).

## RESULTS

### SARS-CoV-2 genome assessment

Illumina sequences obtained using the ARTIC protocol were processed through our customized SARS-CoV-2 genome analysis pipeline (**Fig 1**). This workflow generates consensus genomes and lineage assignments using both Pangolin and Nextclade. Missing data are assessed using customized plots (**Fig 1B**). Missing data plots were used to assess the genomic location of missing data (basecall = N) due to either poor sequence quality or primer dropout. These plots are useful for assessing if a sample failed to amplify (extensive coloring across the plot) or experienced single primer (repeated pattern) or localized (short stretches across samples) dropouts. Consensus genomes were excluded from downstream analysis if they were more than 1,000 bases shorter than the 29,903bp SARS-CoV-2 Wuhan-Hu-1 reference genome. Interactive phylogenetic trees (**Fig 1C**) are also created and visualized on Microreact (https://microreact.org/). In total, we performed ARTIC sequencing on 1,540 samples from which 1,240 consensus genomes passed this size threshold. These consensus genomes were included in all subsequent analyses.

**Figure 1.**
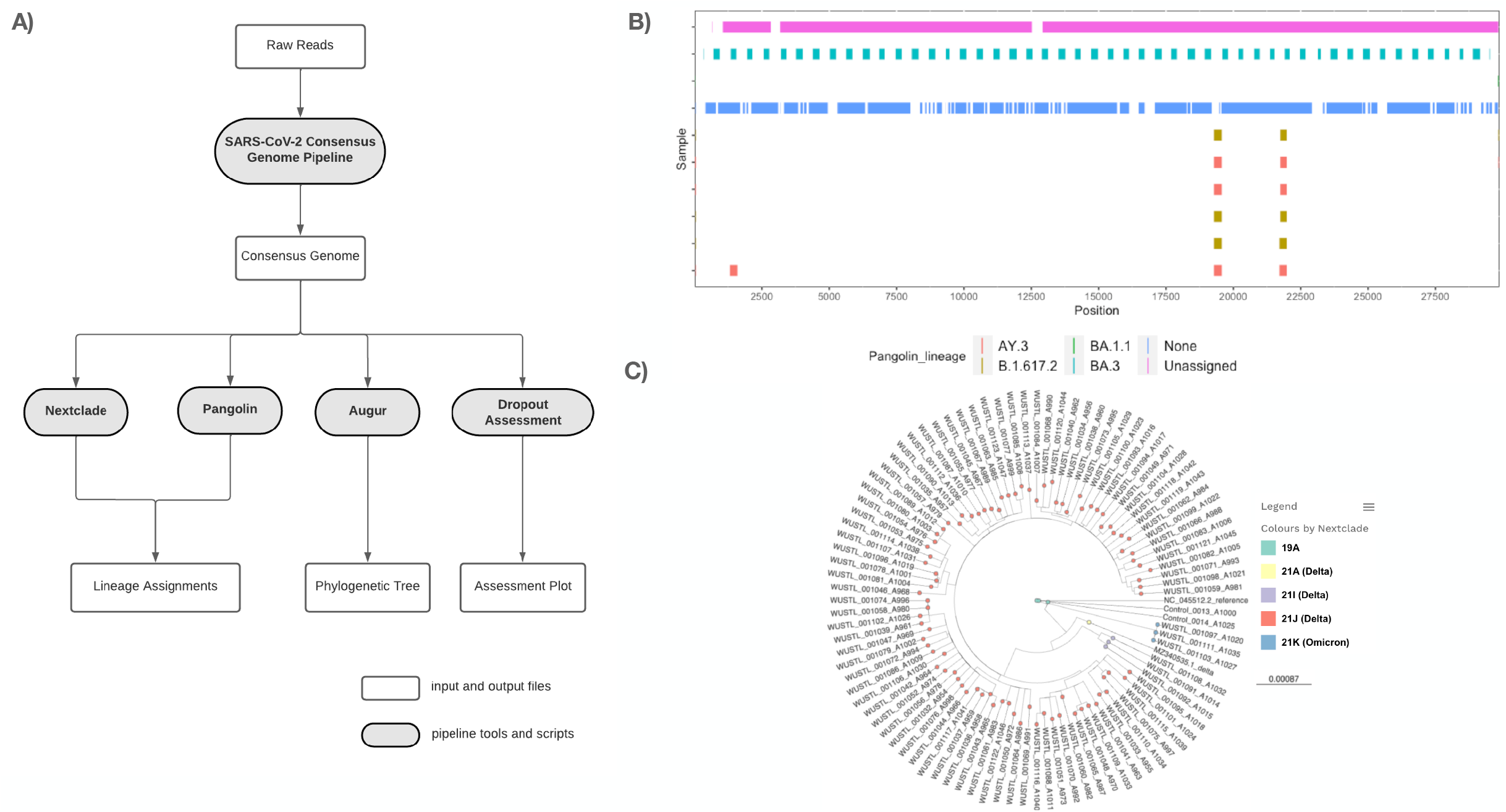
**A)** SARS-CoV-2 genome analysis pipeline. **B)** “Dropout Assessment” Location of missing data (N’s) within a selected subset of SARS-CoV-2 consensus genomes. Colors indicate missing data in specific Pangolin lineage assignments. **C)** Single run phylogenetic tree of SARS-CoV-2 consensus genomes visualized using Microreact.

### Regional versus national VOC prevalence

Consensus genome sequences were classified based on their similarity to known VOC. The proportion of VOC identified biweekly were calculated and compared to national proportions as reported by the Centers for Disease Control and Prevention (CDC) (<u>SARS-CoV-2 Variant Proportions</u>) (**Fig 2**). Regional VOC prevalence exhibited a great deal of parity with national rates. During the earliest phases of the pandemic (Winter through mid-Summer of 2022) genomes with similarity to the Wuhan-Hu-1 reference genome were gradually replaced with strains from the Alpha lineage. During this same period, the Beta lineage of SARS-CoV-2 subtly emerged nationally, but it was not seen in our local genomic surveillance. Delta was first observed nationally during the first 2-weeks of May 2021 but regional detection was slightly delayed until the last 2-weeks. Other than a limited amount of Alpha lineage detection, the Delta lineage had completely taken over both regionally and nationally by mid-July 2021. Omicron was detected both regionally and nationally in early December and completely replaced the Delta variant by the end of January, 2022. Omicron remained the dominant VOC throughout the Spring and Summer of 2022.

**Figure 2.**
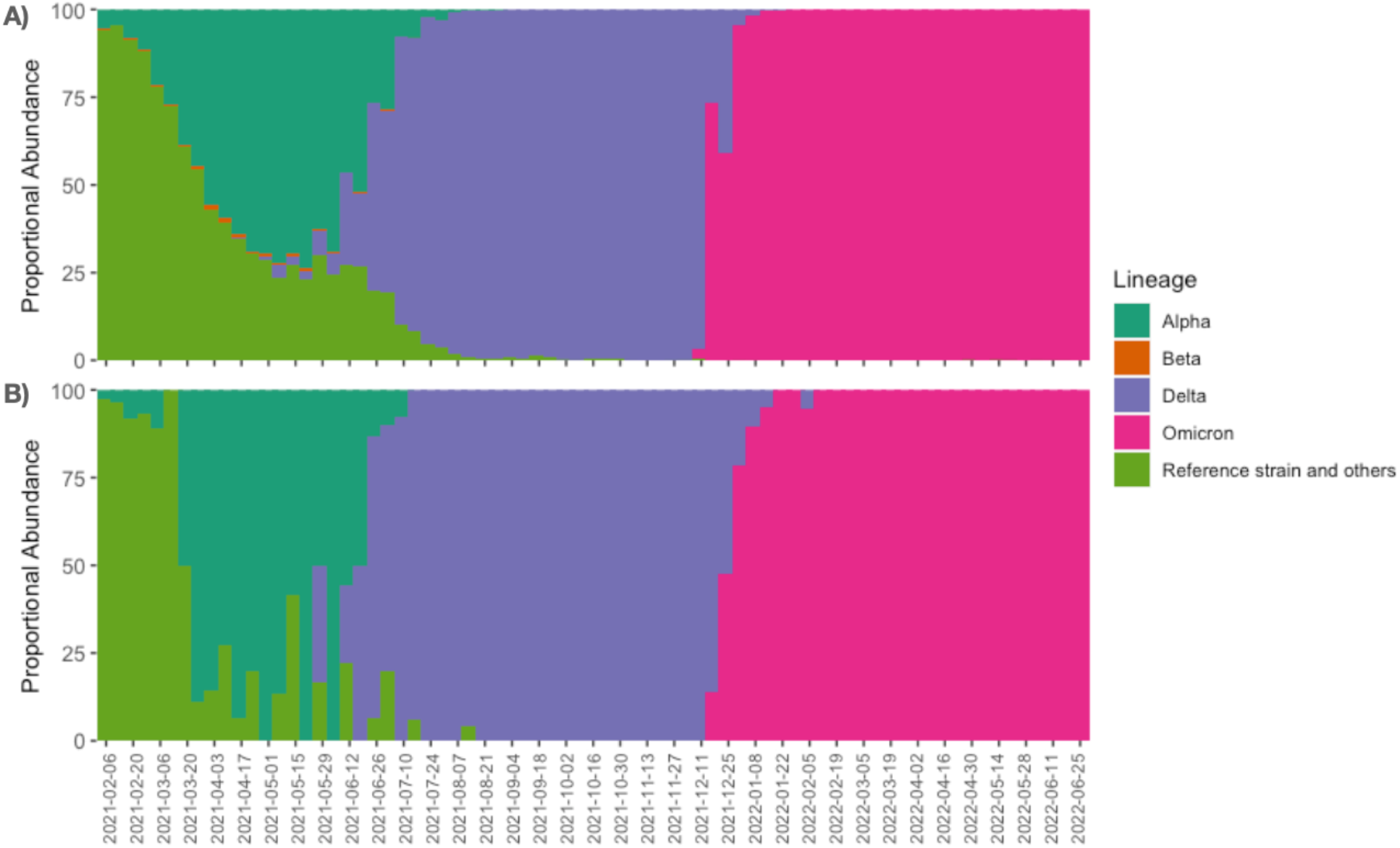
**A)** National VOC prevalence as reported by the Centers for Disease Control and Prevention. **B)** Regional VOC prevalence as detected in the current study.

### Monitoring regional SARS-CoV-2 subvariant lineages

SARS-CoV-2 VOC lineages are composed of a collection of subvariant lineages. In particular, early pandemic spread of the Omicron lineage was characterized by two primary subvariant lineages. Omicron subvariant lineage BA.1 dominated 2021, with subvariant BA.2 emerging during the Winter of 2021/2022 [7]. Our regional genomic surveillance identified similar patterns (**Fig 3**). The only local Omicron subvariants identified between December, 2021 and February 2022 belonged to the BA.1 lineage (BA1, BA1.1, BA.1.1.10, BA.1.1.4 and BA.1.15). Omicron subvariant BA.2 was first detected in February 2022 completely replacing the BA.1 subvariant lineage by the end of April, 2022. BA.2 was dominant throughout the Summer of 2022 with the dominant subvariants belonging to BA.2 and BA.2.12.1. In total 17 BA.2 subvariants were detected throughout this time period. In addition, Omicron subvariant lineage BA.2 and BA.5 began to be detected in April/May 2022 with increasing detection of BA.5.5 through June.

**Figure 3.**
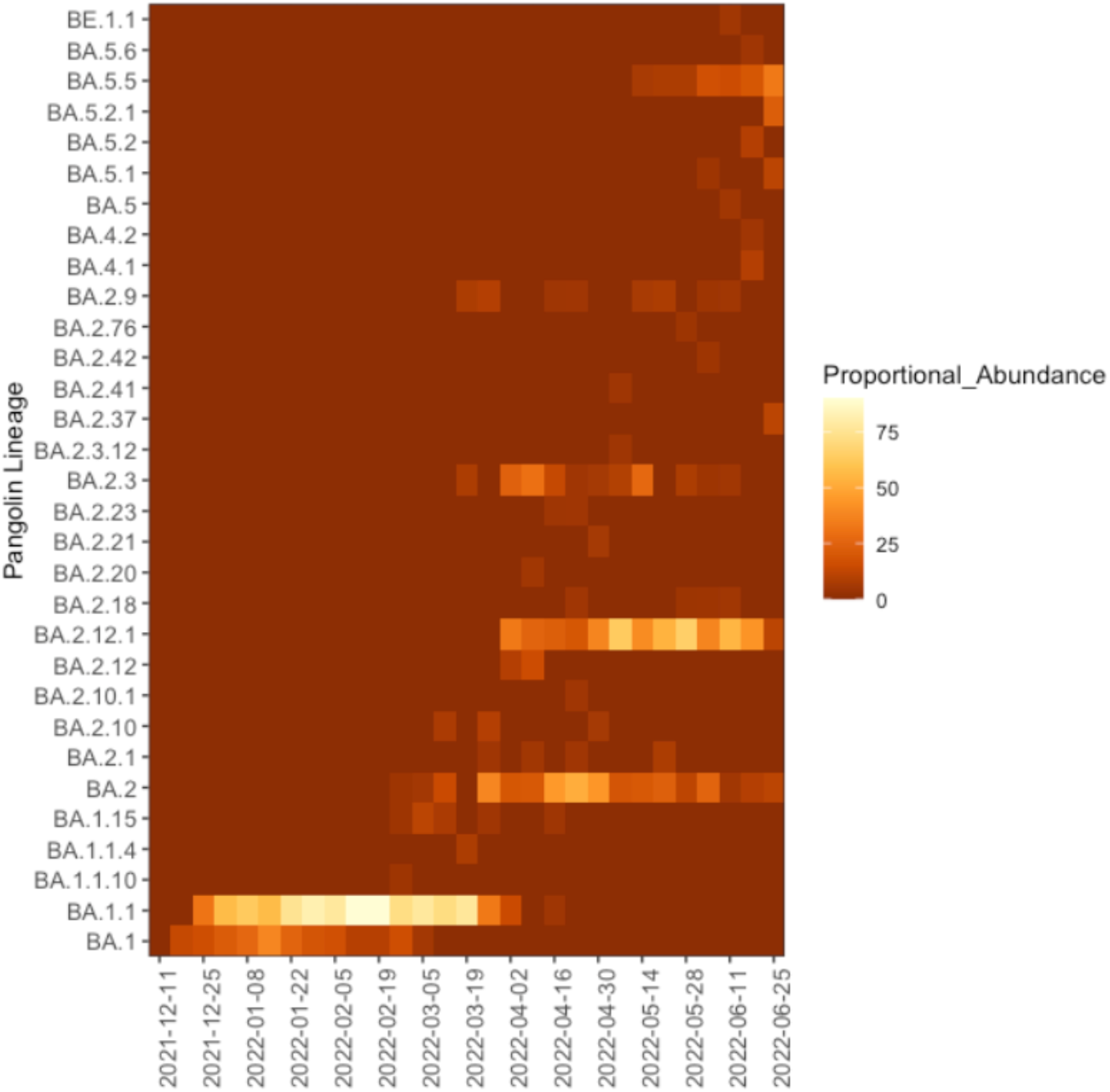
Detection of local SARS-CoV-2 Omicron subvariant lineages between December 2021 and June 2022.

### Analysis of regional genomic sequence divergence from known global references

Sequence divergence from all locally acquired consensus genomes and selected reference genomes were calculated using Nextclade (**Fig. 4**) [13]. All variants were calculated descendants of previously known reference variants. All local consensus genomes clustering within the Alpha or reference-like clades were classified as descendants to B.1.2, B.1.1.7, BA.1.1.519 and several other B.1.X and B.1.1.X sublineages (**Fig. 4A**). All local consensus genomes assigned to the Delta and Omicron VOC were assigned to the 21A or 21J (**Fig. 4B**) and BA.1, BA.2, BA.4 and BA.5 (**Fig. 4C**) subvariant lineages respectively. No local consensus genomes clustered with WHO reference recombinant genomes.

**Figure 4.**
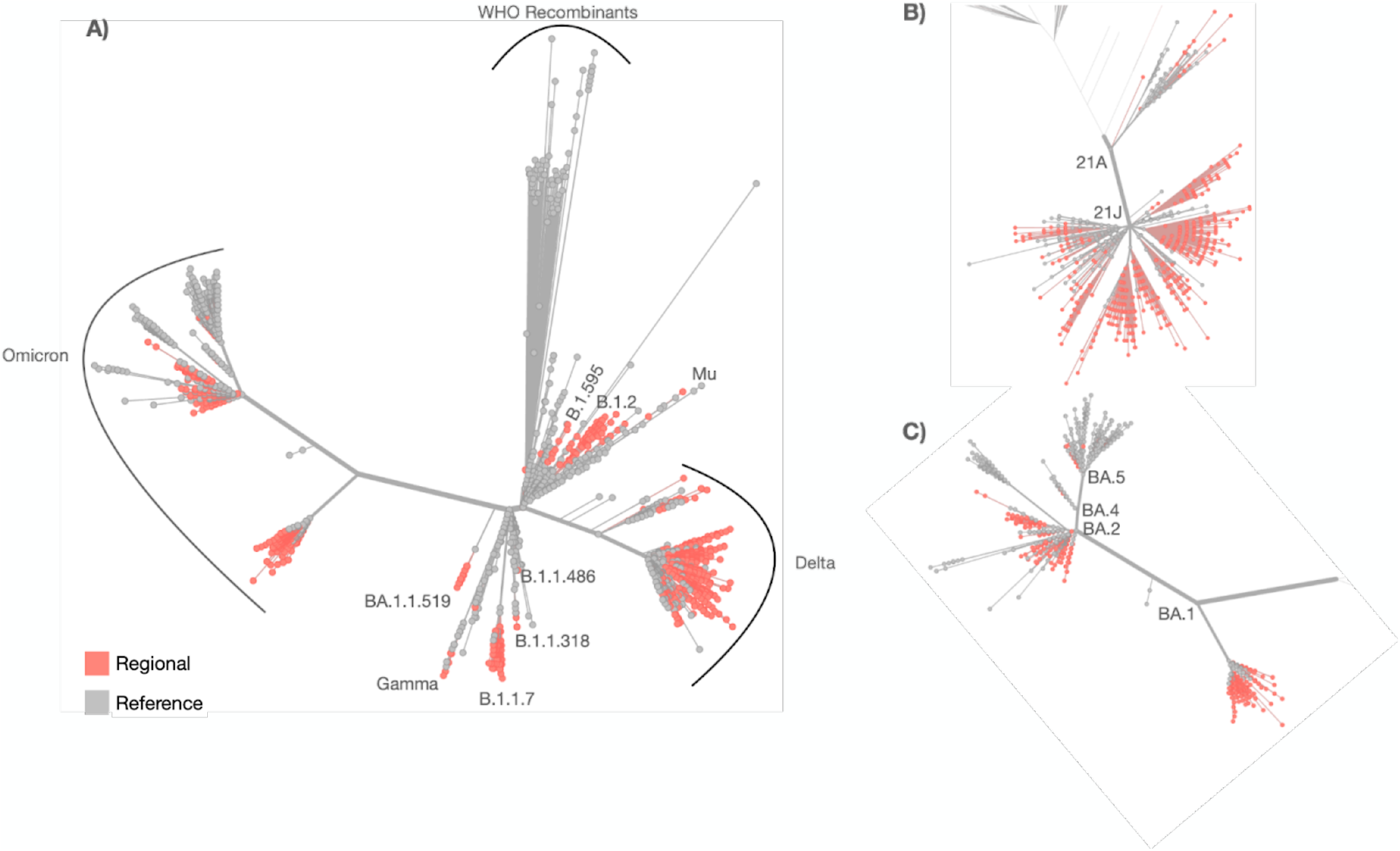
**A)** Phylogenetic tree representing regional sequence divergence in relation to global reference VOC reference genomes. Zoomed in representation of local sequence variants in relation to global reference genomes for **B)** Delta and **C)** Omicron lineages.

### Analysis of deletions and insertions in regional SARS-CoV-2 genomes

We assessed the prevalence and length of deletions and insertions in all local consensus genomes relative to the SARS-CoV-2 Wuhan-Hu-1 reference genome. We identified a large number of genomic deletions of various frequencies and lengths (**FIg 5, Supplemental Table 2**). High-frequency deletions (> 75% frequency) were identified in all lineages of local consensus genomes other than those assigned to the reference strain (Wuhan-Hu-1) lineage where deletion frequency was relatively low (**Fig 5A**). The number of deletions per lineage varied with genomes assigned to the reference strain (n = 17), Alpha (n = 8), Delta (n=47) and Omicron lineages (21K = 15 and 21L = 19) (**Fig 5**). High-frequency (> 75% frequency) deletions occurred most frequently in ORF1ab and the S gene in genomes assigned to the Alpha, Delta and Omicron lineages. Lineage specific high-frequency deletions were identified in the N gene of Alpha and Omicron (21K and 21L), ORF8 of Delta and the 3’ UTR of Omicron 21L. The minimum deletion length across all consensus genomes was 1 base with a maximum deletion length of 126 bases in the Delta lineage within Orf3a (**Fig 5B**). We identified a cluster of deletions within Orf7a of the Delta lineage (**Fig 5B)**. These Delta lineage specific Orf7a deletions had an average start at position 27,588 (min = 27,520, max = 27,721) and an average end at position 27,631 (min = 27,577, max = 27,741) yielding deletions ranging from 1 to 124 bases in length with an average length of 43.5 bases. Each Orf7a deletion was only found in one or two out of 490 local consensus SARS-CoV-2 Delta genomes (lineage frequency range = 0.002% - 0.004%) with the largest 124 base deletion only detected once.

**Figure 5.**
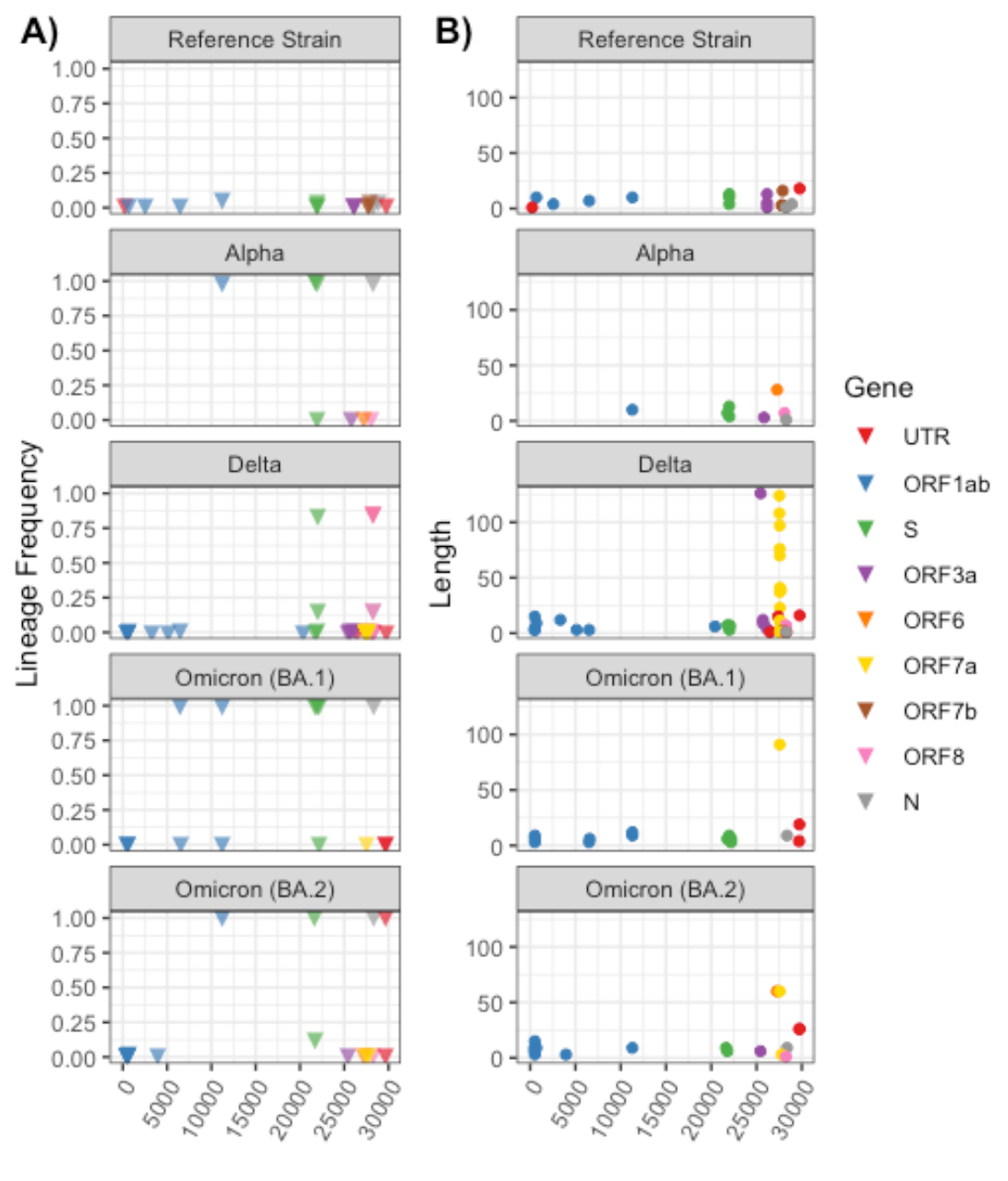
**A)** Lineage specific deletion frequency and **B)** deletion length in local consensus genomes.

Relative to deletions, far fewer insertions were identified in local consensus genomes (**Fig 6, Supplemental Table 2**). There were no insertions found in consensus genomes assigned to the Alpha lineage. Insertions in the S gene were found in consensus genomes assigned to the reference strain and Omicron strains, but were absent for genomes assigned to the Delta lineage. Insertions ranged in length from 1 to 12 bases, with the longest insertion occurring in the S gene of a single Omicron (BA.2) consensus genome.

**Figure 6.**
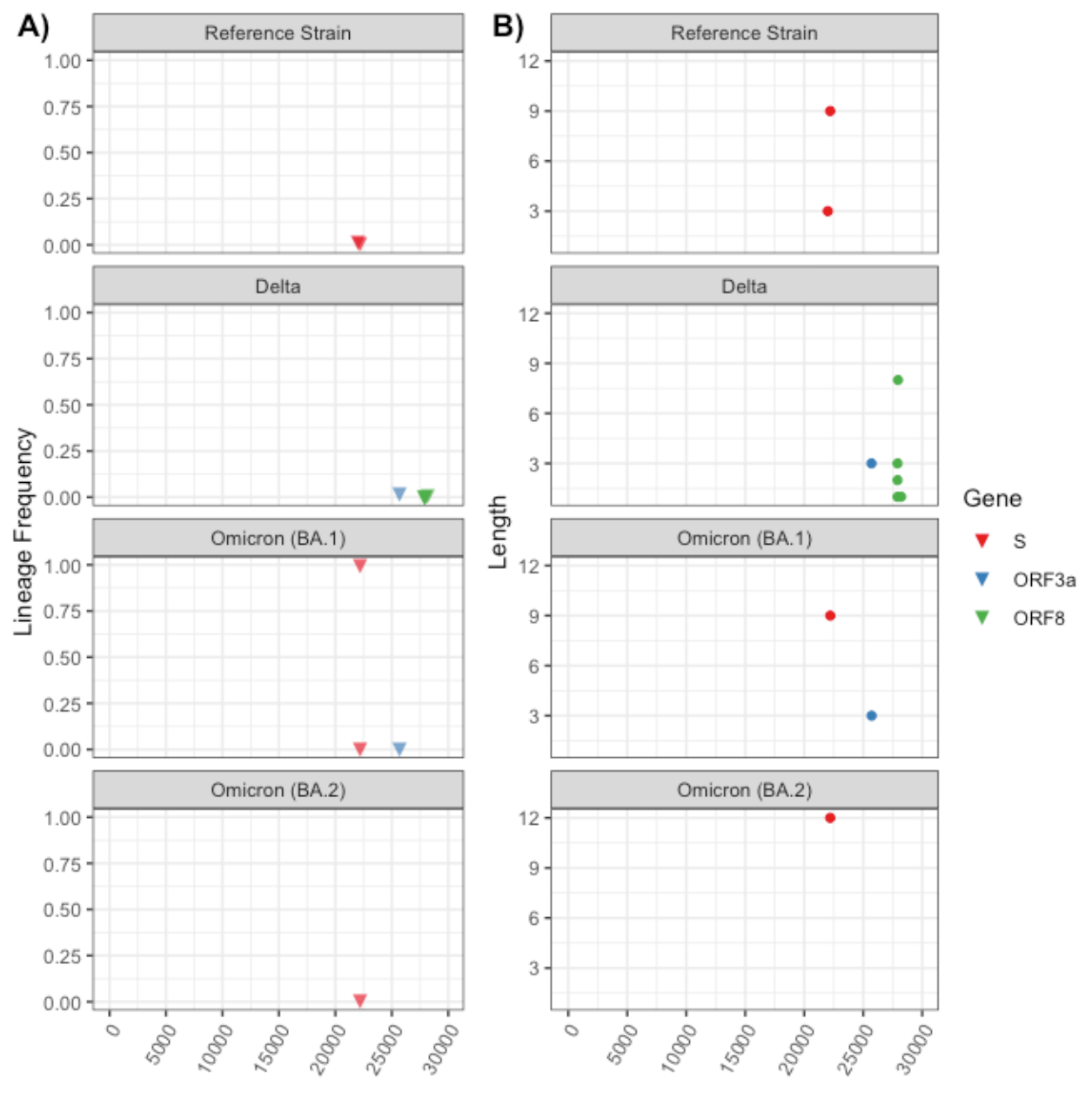
**A)** Lineage specific insertion frequency and **B)** length in local consensus genomes.

## DISCUSSION

Pandemics are a global phenomenon comprising a network of local, community driven outbreaks. Understanding global spread starts at the local level. Conversely, understanding how local infection dynamics compare to global trends can provide regional information about the emergence and circulation of globally identified strains. In addition, genomic surveillance can provide highly-detailed genetic maps of regionally circulating strains.

Building a local genomic surveillance program requires access to the appropriate infrastructure including a patient-facing diagnostic lab for detection and procurement of samples containing the pathogen of interest, a molecular biology lab, modern sequencing infrastructure and bioinformaticians for data processing and analysis. Availability to these resources will vary from region-to-region, but a universal consideration will be sampling depth and cadence. Ultimately, sampling will depend on the goals of the genomic surveillance program as well as financial considerations and laboratory throughput. While access to resources and financing varies across regions, it is still useful to place some boundaries for building a successful sampling framework.

Efforts have been made to model the sampling required to detect the prevalence and emergence of SARS-CoV-2 VOC using regional genomic surveys [16]. These models estimate that VOC detection and average prevalence can be sufficiently measured sequencing ∼10 samples/1,000,000 people/week. While models are important guides for designing sampling strategies, real world efforts can be useful for model calibration and validation. In the present study we assessed our ability to detect globally circulating SARS-CoV-2 VOC in a metropolitan hospital in St. Louis, MO, USA. We sampled ∼5 samples/1,000,000 people per week (n = 1,240 samples), half of the requirements of the predicted model. We detected all significant SARS-CoV-2 VOC (Alpha, Delta and Omicron) and were also able to detect sublineage variation circulating within the community. Our data support that low number consistent sampling is sufficient to detect the prevalence of known and novel VOC within a community.

In addition to determining local VOC prevalence, regional genomic surveillance provides a wealth of detailed genetic information. While low-frequency subvariants may only appear transiently in a region and never achieve global spread, they still contain genomic variation useful for mechanistic studies. For example, a 115 base pair deletion in SARS-CoV-2 ORF7a (27,549–27,644 nt) was identified as part of regional genomic sequencing efforts in Bozeman, Montana, USA [17]. This ORF7a deletion (ORF7a^Δ115^) was subsequently shown to have an *in vitro* growth defect associated with elevated IFN response suggesting an immunosuppressive role for ORF7a. In our study, we also identified a collection of deletions in the Orf7a gene in strains from the Delta lineage. Deletions in Orf7a were uncommon, but were as large as 124 bases long. These Orf7a and other subvariant isolates can serve as a natural laboratory bridging real world genetic variation with detailed mechanistic studies.

Taken together, we demonstrate that modest sampling efforts paired with robust infrastructure can provide genomic surveillance of SARS-CoV-2 at a regional level. These efforts can characterize the presence and prevalence of VOC. In addition, regional genomic surveillance generates a repository of unique genetic variation. We believe these results should serve as a guideline for future SARS-CoV-2 genomic surveillance programs.

## Data Availability

All whole viral genomes were made publicly available at GenBank.

## Author Contributions

B.A.P and D.W. conceptualized the study. B.A.P. coordinated all sample collection. L.D., C.F. and L.C. organized sample extraction and sequencing A.J, B.F. and S.A.H. completed all bioinformatic analysis, and figure design and creation. Writing was shared evenly across all authors.

